# Shared Polygenic Architecture Across Arteriopathies: An Integrative Cross-Trait Analysis

**DOI:** 10.64898/2026.06.18.26356018

**Authors:** Stephen O. Brennan, Alexander C. Tinworth, Iyas Daghlas, Quentin Le Grand, Bastien Rioux, Peter Kelly, Dipender Gill, Stéphanie Debette, John J. McCabe, the CADISP Consortium

**Affiliations:** Health Research Board (HRB) Stroke Clinical Trials Network Ireland (SCTNI), Ireland; School of Medicine, University of Galway, Ireland; National Institute for Prevention and Cardiovascular Health, Galway, Ireland; Nuffield Department of Population Health, University of Oxford, Oxford, UK; Department of Neurology, UCSF Weill Institute for Neurosciences, University of California San Francisco; University of Bordeaux, INSERM, Bordeaux Population Health research center, UMR1219, F-33000, Bordeaux, France; Institut du Cerveau (ICM), Paris Brain Institute, INSERM U1127, UMR CNRS 7225 Paris, Sorbonne Université, Assistance Publique des Hôpitaux de Paris, Paris, France; Department of Neurology, Mater Misericordiae University Hospital, Dublin, Ireland; School of Medicine, University College Dublin, Dublin, Ireland; Department of Epidemiology and Biostatistics, School of Public Health, Imperial College London, London, UK; Stroke Service, Department of Geriatric Medicine, Mater Misericordiae University Hospital, Dublin, Ireland

**Keywords:** genetic correlation, aortic aneurysm, cervical artery dissection, spontaneous coronary artery dissection, intracranial aneurysm, Mendelian randomization

## Abstract

**Background:** Non-monogenic arteriopathies are often classified as distinct entities according to the arterial territory involved, yet they share clinical features and may co-occur in the same individual. This pattern suggests shared susceptibility across anatomically distinct arteriopathies, potentially driven by common biological and genetic mechanisms.

**Methods:** We investigated the shared genetic architecture of five arteriopathies (cervical artery dissection (CeAD), intracranial aneurysm (IA), spontaneous coronary artery dissection (SCAD), aortic aneurysm and dissection (AAD), and fibromuscular dysplasia (FMD)) using LD score regression, Association analysis based on SubSETs (ASSET), pairwise Multi-Trait Analysis of Genome-wide association summary statistics (MTAG), pleiotropy mapping and Mendelian randomization (MR) to identify shared loci and prioritise candidate causal genes.

**Results:** LD score regression identified significant positive genetic correlations between CeAD-SCAD (rg = 0.64), IA-AAD (rg = 0.33), IA-SCAD (rg = 0.37), CeAD-AAD (rg = 0.56) and SCAD-AAD (rg = 0.20). ASSET identified 37 shared independent loci, and in MTAG analyses, one novel locus was identified for CeAD and SCAD (*SLC39A8*) and one for IA (*FGF5*). 13 loci showed strong cross-trait colocalization, including *PHACTR1*, *LRP1*, and *CDKN2B-AS1*.

Using the Genotype-Phenotype Map, we found that arteriopathy-associated variants colocalized with blood pressure-and migraine-related traits, while many showed effect directions opposite to those observed for coronary artery disease. Proteome-wide MR identified 67 circulating proteins associated with at least one trait, including ECM1 and SHISA5 for CeAD and FGF5 for IA, with 17 supported by colocalization. Transcriptome-wide MR identified 204 colocalized tissue–specific signals, of which, 14 were shared across multiple traits. Enrichment analyses implicated pathways related to vascular development, smooth muscle cell function, extracellular matrix organization, and TGF-β signaling.

**Conclusions:** These findings support shared genetic architecture across anatomically distinct arteriopathies, implicating pathways involved in vascular structure and prioritising therapeutic targets for future mechanistic investigation.

## Introduction

Rare monogenic arteriopathies such as Marfan Syndrome and Ehlers-Danlos syndrome type IV are typically caused by monogenic mutations affecting extracellular matrix components or TGF-β signaling pathways and are often identifiable through clinical genetic testing.^1,2^ In contrast, arteriopathies such as intracranial aneurysm (IA), aortic aneurysm and dissection (AAD), fibromuscular dysplasia (FMD), spontaneous coronary artery dissection (SCAD), and cervical artery dissection (CeAD), have more complex, incompletely characterized genetic architectures, with pathogenic variants in known monogenic disease genes identified in a minority of cases.^3,4^ Despite involving distinct arterial territories, these conditions share common clinical features, including relatively early onset, limited contribution of traditional atherosclerotic risk factors, and, for several, a marked female predominance.^4–6^

These arteriopathies also show evidence of clustering within individuals. SCAD is often accompanied by extracoronary dissections or aneurysms, and similar multi-territory involvement is seen in CeAD and IA, even in the absence of monogenic connective tissue disorders.^7–10^ Furthermore, FMD, a non-atherosclerotic medium-vessel arteriopathy, is over represented across these conditions.^10,11^ Taken together, this aggregation of anatomically distinct arteriopathies within individuals suggests shared underlying susceptibility, yet the genetic basis for this convergence remains largely unexplored.

Here we investigate the shared genetic architecture of arteriopathies using integrative cross-trait analyses. We show that these traits share genetic liability, identify pleiotropic loci, and find that blood pressure-related traits are prominent among colocalizing phenotypes, while many arteriopathy-associated variants show opposite directional effects to those for coronary artery disease (CAD). Finally, using Mendelian randomization (MR), we prioritize genes implicated in individual arteriopathies. Identifying shared loci, genes, and biology may improve risk stratification, diagnosis, and therapeutic development across the spectrum of arteriopathies.

## Methods

### Study population

Summary statistics were obtained from published, European ancestry consortium-based genome-wide association studies (GWAS) of SCAD (1,917 cases; 9,292 controls),^12^ CeAD (1,393 cases; 14,416 controls),^13^ FMD (1,556 cases; 7,100 controls),^14^ IA (7,495 cases; 71,934 controls)^15^ and AAD (22,049 cases; 419,651 controls).^16^ Genotypic quality control was performed on each GWAS by harmonizing summary statistics to the 1000 Genomes Project European reference panel (GRCh37). All contributing studies obtained appropriate participant consent and received approval from the relevant ethics committees or institutional review boards. Supplementary Table 1 lists basic characteristics of all included GWAS and Figure 1 outlines the study’s overall design.

**Figure 1.**
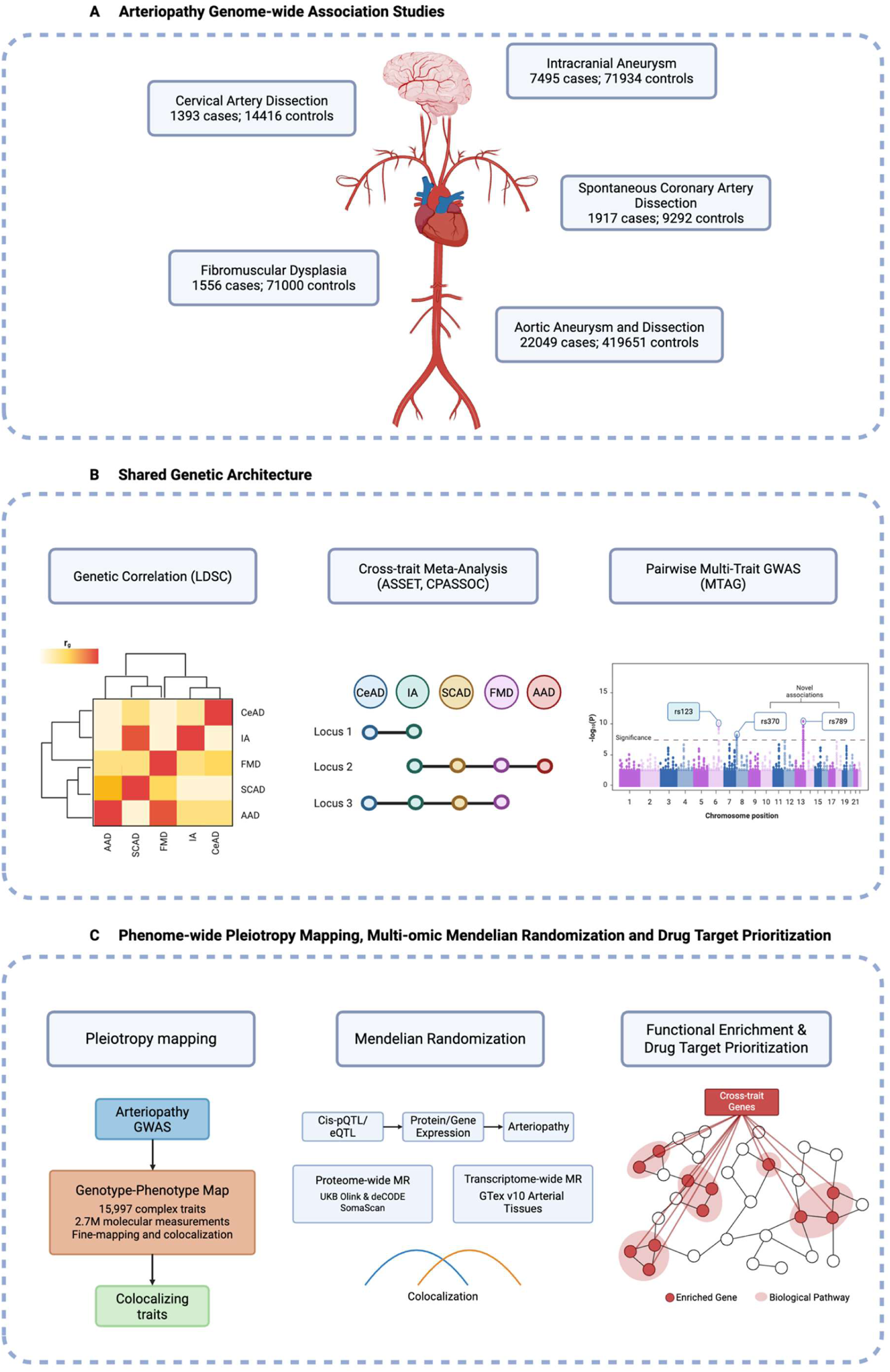
Study design for integrative genetic analyses across arteriopathies Created in BioRender. Brennan, S. (2026) https://BioRender.com/ow2ilk5

### LD score regression

To estimate SNP-based heritability (h²ₛₙₚ) and pairwise genetic correlations, we used linkage disequilibrium (LD) Score Regression (LDSC v2.0.0).^17^ Genetic covariance intercepts were used to assess sample overlap between studies. Pairwise genetic correlations (n=10) were corrected for multiple testing using the Benjamini-Hochberg false discovery rate (FDR) method.

### Cross-trait association analysis

To systematically investigate shared and heterogeneous genetic effects across arteriopathies, we applied three complementary cross-trait approaches: (1) Association analysis based on SubSETs (ASSET)^18^, which tests each genetic variant for association across traits and identifies the subset contributing to the strongest joint signal; (2) Cross-phenotype association analysis using summary statistics from GWAS (CPASSOC)^19^, which accommodates heterogeneous effects across traits; and (3) Multi-Trait Analysis of GWAS (MTAG)^20^, which leverages genetic correlations to improve trait-specific power to detect genetic associations. Further details on these methods are provided in the Supplementary Methods.

### Fine-mapping and trait–trait colocalization analysis

Lead loci identified through ASSET were fine-mapped within each contributing trait using Sum of Single Effects regression (SuSiE-RSS) (±500 kb windows, and LD matrices from 337,491 unrelated British-ancestry UK Biobank participants).^21^ Ridge regularization (λ = 10⁻³) was applied to LD matrices for numerical stability.

For ASSET-identified pleiotropic loci, cross-trait colocalization was performed between all pairwise combinations of contributing traits to identify shared causal variants underlying multi-trait associations. Colocalization was assessed using coloc.susie with default priors (p1 = p2 = 10⁻⁴, p12 = 10⁻⁵). At loci where SuSiE failed to converge, coloc.abf was applied. Colocalization posterior probability for a shared causal variant (PP.H4) was classified as strong (≥0.80), moderate (0.50–0.80), or weak (<0.50).

### Functional mapping and annotation

Lead signals (and fine-mapped variants where available) were mapped to candidate causal genes using a hierarchical evidence framework prioritizing the most direct functional support. Protein-altering variants were assigned to genes based on predicted consequence using Ensembl Variant Effect Predictor. Non-coding variants were mapped to genes with arterial tissue eQTL signals that colocalized with the GWAS association (coloc, PP.H4 > 0.8) in GTEx v10 (aorta, coronary artery, tibial artery). Variants with regulatory evidence were additionally mapped using enhancer-to-gene (E2G) scores from Open Targets. Variants lacking functional or regulatory support were assigned to the nearest protein-coding gene.

### Novel loci definition

To identify novel associations, MTAG lead variants were compared against previously reported associations in the GWAS Catalog. A locus was considered novel if the lead variant was more than 500 kb from, and not in linkage disequilibrium (r² < 0.1) with, any variant previously associated with the same trait.

### Genotype-Phenotype Map

To characterize the shared phenotypic architecture of arteriopathy-associated loci, we interrogated each univariate GWAS against the Genotype-Phenotype Map (GPMap), a repository of colocalizing genetic associations across 15,997 complex traits and 2.7 million molecular measurements. The GPMap pipeline performs summary statistic imputation, SuSiE fine-mapping, pairwise colocalization, and graph-based clustering of shared signals into colocalization groups. We applied a regional significance threshold of *P* < 1 × 10^-5^ for variant extraction. Full details of the GPMap framework are described elsewhere.^22^ To assess whether arteriopathy-associated alleles showed concordant or discordant effects with CAD, we extracted lead variants from all loci identified by GPMap colocalization and obtained effect estimates from the corresponding arteriopathy summary statistics and a GWAS of CAD.^23^ Effects were aligned to the CAD risk-increasing allele and loci reaching genome-wide significance (*P* < 5×10⁻⁸) in both the arteriopathy and CAD were classified as concordant or discordant.

### Proteome-wide Mendelian randomization

To investigate the effect of circulating protein abundance on arteriopathies, we performed two-sample MR using cis-pQTLs obtained from published proteomic GWAS in the UK Biobank Pharma Proteomics Project (Olink Explore 3072; 2,923 proteins measured in 34,557 participants)^24^ and the deCODE Health Study (SomaScan v4; 4,719 proteins measured in 35,559 participants).^25^ Instruments were selected using the original study-wide significance thresholds (*P* < 1.7 × 10⁻¹¹ for Olink; *P* < 1.8 × 10⁻⁹ for SomaScan), and were restricted to variants within ±100 kb of the encoding gene. Where multiple cis-pQTLs were available, variants were clumped at r² < 0.001 using the 1000 Genomes Project European reference panel. Instruments with *F*-statistic < 10 were excluded.

To replicate the causal estimates from the proteome-wide MR, we performed two-sample cis-MR using cis-pQTL instruments from previously published proteomic GWAS carried out in different cohorts, including SCALLOP (Olink; n = 30,931),^26^ and the Fenland Study (SomaScan; n = 10,708) where available.^27^ The same instrument selection and harmonization procedures as in the primary analyses were applied. Associations showing Bonferroni-corrected significance (*P* < 0.05) and with concordant direction of effect were considered replicated.

### Transcriptome-wide Mendelian randomization

To investigate the effect of gene expression on arteriopathies, we performed two-sample MR using cis-expression quantitative trait loci (cis-eQTLs) from the Genotype-Tissue Expression (GTEx) Consortium v10.^28^ Genome-wide significant (*P* < 5 × 10⁻⁸), uncorrelated (r² < 0.001) variants located within ±100 kb of each gene were selected from European ancestry samples across aortic, coronary artery, tibial artery, whole blood tissues and fibroblasts. Variant harmonization and clumping followed the same procedures as described for proteome-wide MR.

### Mendelian randomization analyses

MR estimates were obtained using the Wald ratio for single-variant instruments and inverse-variance weighted models when multiple independent instruments were available.

Associations were adjusted for multiple testing using the Storey false discovery rate (q-value) method, with an FDR threshold of 5% applied separately for each outcome trait. The same framework was additionally applied to systolic blood pressure, diastolic blood pressure and pulse pressure using genome-wide significant instruments from UKB^29^, clumped at *r*^2^ < 0.001 within a 10-Mb window.

### Colocalization

For proteins or genes with significant MR associations, colocalization was performed to assess whether the QTL and disease signals shared a causal variant. We applied coloc.susie to ±500 kb regions centered on lead variants, using SuSiE-derived credible sets and default priors (p1 = p2 = 10⁻⁴, p12 = 10⁻⁵). At loci where SuSiE failed to converge, coloc.abf was applied. Posterior probability of a shared causal variant (PP.H4) ≥ 0.80 was considered strong evidence of colocalization, while (PP.H4) 0.50–0.80 was considered suggestive.

### Functional enrichment analyses

Gene-set enrichment was performed using MAGMA on ASSET summary statistics. Tissue expression and cellular component enrichment of prioritized genes were assessed using FUMA GENE2FUNC with GTEx v8 and Gene Ontology Cellular Component annotations.^30^ Genes from FDR-significant pQTL and eQTL MR associations were pooled across outcomes and analysed using clusterProfiler. Trait-specific enrichment was performed as a sensitivity analysis, and pathway convergence was assessed across MAGMA, pQTL-MR, and eQTL-MR results.

### Drug target assessment

Identified genes were queried against the Open Targets Platform (v24.12) via the GraphQL API to assess tractability and identify existing pharmacological agents. Tractability assessments encompassed small molecule (SM), antibody (AB), proteolysis targeting chimeras (PR), and other clinical (OC) modalities. “Known drugs” were defined as compounds with approved or investigational indications linked by Open Targets to the gene or pathway.

### Data availability

GWAS summary statistics for IA, AAD, FMD, and SCAD are publicly available through the GWAS Catalog (https://www.ebi.ac.uk/gwas/). Summary statistics for CeAD are available upon request to the CADISP consortium investigators. Cis-pQTL data were obtained from the UK Biobank Pharma Proteomics Project (available via Synapse, https://metabolomics.helmholtz-munich.de/ukbbpgwas/) and the deCODE Health Study (https://www.decode.com/summarydata/). QTL data were obtained from GTEx v10 (https://gtexportal.org/). Analyses were conducted using R (v4.3.3) and Python (v3.12.11). The analysis code used to generate the results and figures in this study will be made available at time of publication.

## Results

After filtering to HapMap3 SNPs, between 0.39 (IA) and 1.17 million variants (AAD) remained for analysis. SNP-based heritability (h²SNP) on the observed scale ranged from 0.02 (AAD) to 0.29 (IA) (Table S2).

### LD score regression

Pairwise genetic correlations were estimated using LD score regression (Figure 2; Table S3). Five of ten trait pairs showed significant positive genetic correlation after FDR correction. The strongest correlation was observed between CeAD and SCAD (rg = 0.64, *P*_fdr_ = 0.048). IA was significantly correlated with AAD (rg = 0.33, *P*_fdr_ = 0.009) and SCAD (rg = 0.37, *P*_fdr_ = 0.033). AAD was also correlated with CeAD (rg = 0.56, *P*_fdr_ = 0.048) and SCAD (rg = 0.20, *P*_fdr_ = 0.048).

**Figure 2.**
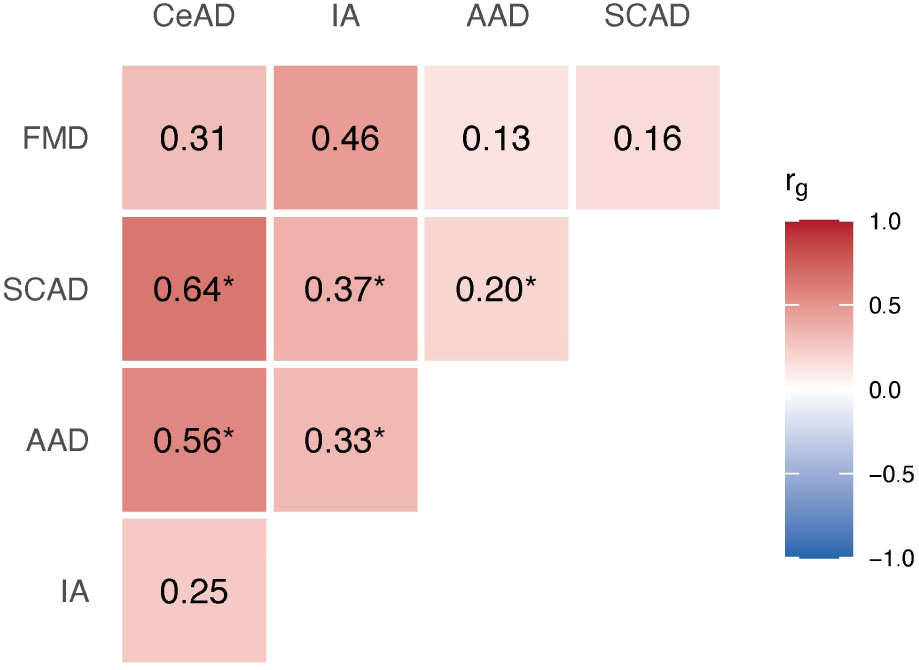
Pairwise LD score regression across five arteriopathy traits. *denotes false discovery rate *P* <0.05. AAD indicates aortic aneurysm; SCAD, spontaneous coronary artery dissection; IA, intracranial aneurysm; FMD, fibromuscular dysplasia; CeAD, cervical artery dissection.

### Cross-trait association analysis

Using ASSET, we identified 37 independent genome-wide significant loci showing cross-trait effects across arteriopathies, with each contributing trait in the optimal subset showing at least nominal univariate support (P < 0.05) (Table S4). Twenty-seven loci (73%) showed concordant effect directions across contributing traits. The strongest concordant signal across all five traits mapped to 9p21 locus (*CDKN2B-AS1*; *P* = 8.3 × 10⁻⁵⁹). Other concordant loci contributing to four or more traits included *LRP1* (*P* = 8.1 × 10⁻⁴³; CeAD, FMD, SCAD, AAD) and *ECM1* (*P* = 5.6 × 10⁻¹⁹; CeAD, FMD, SCAD, AAD). Ten loci (27%) exhibited effects in opposite directions across contributing traits. At *MRPS6*, the risk allele for IA was protective for extracranial arteriopathies.

We assigned 35 candidate genes to ASSET loci using a hierarchical evidence framework that included SuSiE fine-mapped credible sets where available. Gene prioritization was supported for 25 loci (71%), based on arterial eQTL colocalization (19 loci; GTEx v10), regulatory chromatin interaction evidence (2 loci), a protein-altering variant in *STARD13*, convergent multi-omic support for ECM1, and prior functional evidence implicating *CDKN2B-AS1* (Supplementary Table 5). At the 6q25.3 locus (nearest gene SLC22A3), fine-mapping resolved the AAD signal to *LPA*. No credible sets were identified for CeAD or FMD, and cross-trait colocalization confirmed distinct causal variants for AAD and FMD (PP.H4 = 0.21). In a complementary cross-trait analysis, CPASSOC (SHet) identified 27 loci, with 26 (96%) mapping to ASSET-identified genes (Table S6).

### Genetic loci discovery through multi-trait analysis of GWAS

To increase power for locus discovery, we applied pairwise MTAG to trait pairs with significant genetic correlations: SCAD-CeAD (rg = 0.64), AAD-CeAD (rg = 0.56), and AAD-IA (rg = 0.33). Estimated maximum false discovery rates ranged from 0.009 to 0.28 (Table S7).

Pairwise MTAG identified 64 genome-wide significant lead variants across four arteriopathies (*P* < 5×10⁻⁸; P < 4.6×10⁻¹¹ for AAD). Of these, 51 showed supporting univariate evidence in the target trait (*P* < 0.001) with concordant effect directions. After LD clumping, these corresponded to 44 unique lead variants across 37 independent loci (Figure 3; Table S8). Seven trait-specific associations (six loci) reached genome-wide significance only through MTAG analysis.

**Figure 3.**
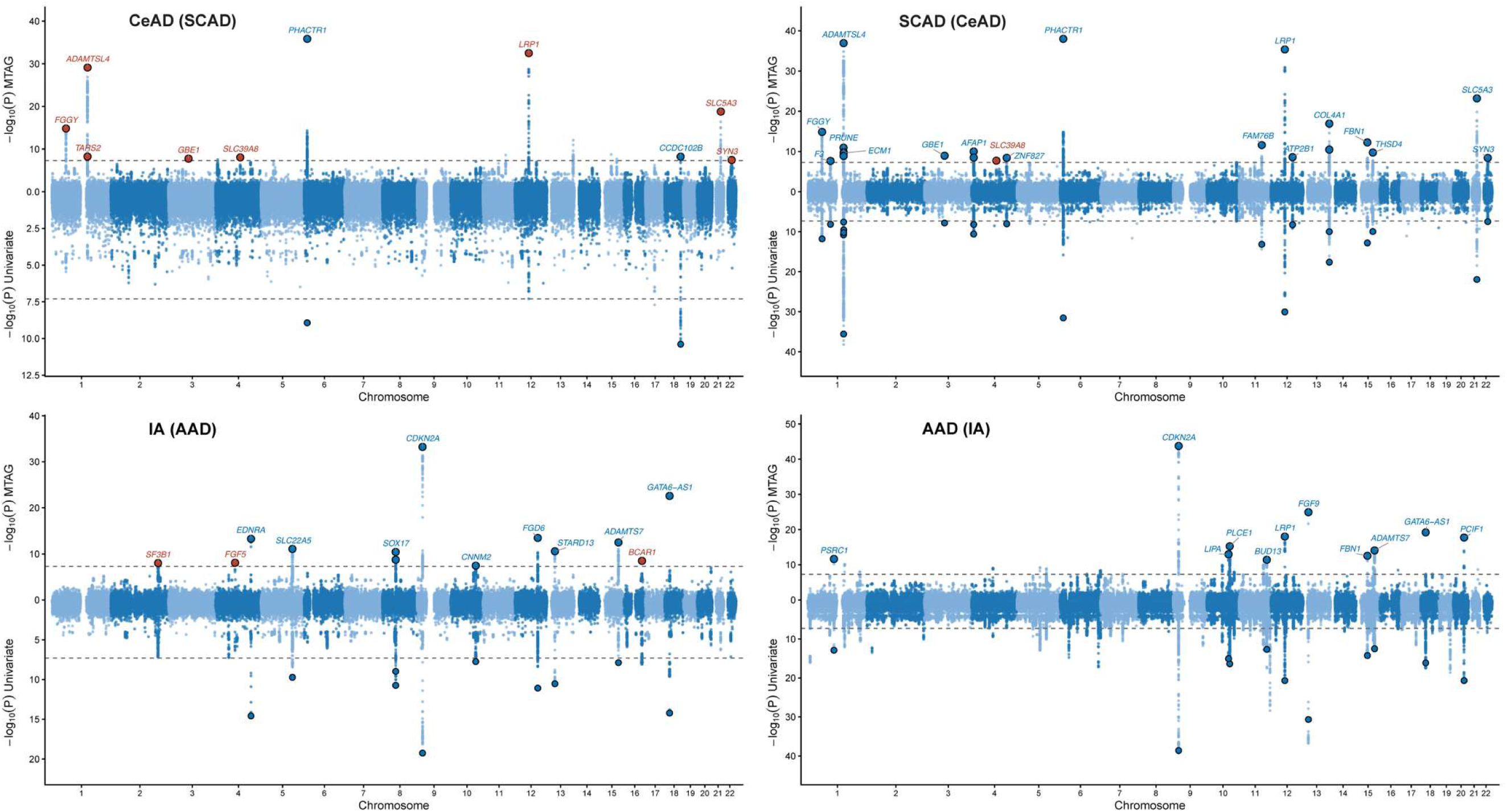
Genetic discovery across five arteriopathy traits. Each panel shows –log₁₀(P) across autosomes for the single-trait GWAS (bottom) and pairwise MTAG (top). Each panel shows MTAG results for the named trait using the partner trait indicated in parentheses. The black dashed line marks genome-wide significance level (*P* < 5 × 10⁻⁸). All loci that only reached significance through MTAG are annotated in red.

Three associations at two loci have not been previously reported in GWAS Catalog for the target arteriopathy: *SLC39A8* (rs13107325), which was novel for both CeAD and SCAD, and *FGF5* (rs12509595), which was novel for IA. Locus-zoom plots for newly identified loci are provided in the Supplementary Material. CeAD showed the greatest increase in association signal after MTAG (mean Δ−log₁₀*P* = 13.0), whereas AAD showed modest attenuation (Δ−log₁₀*P* = −0.8).

The 44 lead variants mapped to 37 unique genes (Table S8). Of these, 26 variants (59%) had functional evidence supporting gene assignment, including 20 with arterial eQTL colocalization, three coding variants, and three supported by chromatin interaction evidence (Table S8). Eight genes were associated with more than one arteriopathy, including *LRP1* (AAD, CeAD, SCAD), *PHACTR1* (CeAD, SCAD), *ADAMTS7* (AAD, IA) and *CDKN2A* (AAD, IA) (Figure 3). Effect directions were concordant across traits, except at *FBN1*, where different lead variants were identified for SCAD (rs7174973) and AAD (rs1036477).

### Cross trait colocalization

To assess whether ASSET loci reflect shared causal or distinct variants, we performed pairwise colocalization. Of 108 pairwise tests across 37 loci, 23 trait pairs showed strong evidence for shared causal variants (PP.H4 ≥ 0.80) and 9 showed moderate evidence (PP.H4 0.50–0.80) (Table S10). The strongest evidence was observed at *PHACTR1* (PP.H4 = 1.0; CeAD, FMD, SCAD), where fine-mapping resolved a single causal variant (rs9349379) with PIP ≈ 1.0 across all three traits, and *LRP1* (PP.H4 > 0.97; CeAD, FMD, SCAD, AAD). At the 9p21 locus (*CDKN2B-AS1*), colocalization supported shared causal variants between FMD and both IA and AAD (PP.H4 > 0.92), but fine-mapping identified distinct causal variants for IA and AAD (PP.H3 ≈ 1.0), consistent with multiple independent signals.

### Pathway enrichment analysis

Gene-based association analysis using MAGMA identified 47 genes at Bonferroni-corrected significance (*P* < 2.8 × 10⁻⁶; Table S11), including *ECM1*, *CDKN2B*, *LRP1*, *MMP3*, and *LOXL1*. Gene-set enrichment revealed convergence on extracellular matrix biology (Extracellular Matrix Disassembly, Elastic Fibre Formation), TGF-β signaling (TGF-β Receptor Binding, Response to TGF-β), and vascular remodeling (Epithelial-to-Mesenchymal Transition) (*P* < 0.05; Table S12). Prioritized genes were enriched among genes upregulated in arterial tissues (Table S13; Figure S3), and showed cellular component enrichment for external encapsulating structure, collagen-containing extracellular matrix and basement membrane (Table S14). Trait-specific enrichment results are provided in the Supplementary Material.

### Genotype-Phenotype Map

To characterize the broader phenotypic profile associated with arteriopathy-associated genetic variation, we used each arteriopathy GWAS as an anchor trait in GPMap, a repository of colocalizing genetic associations across 15,997 complex traits and 2.7 million molecular measurements. Across the five arteriopathies, variants at 58 loci for AAD, 12 for SCAD, eight for IA, four for FMD and two for CeAD colocalized with at least one complex trait (Table S15). Blood pressure-, coronary artery disease-, and migraine-related traits were among the most consistently identified phenotypes across arteriopathies (Figure 4a, Table S15).

**Figure 4.**
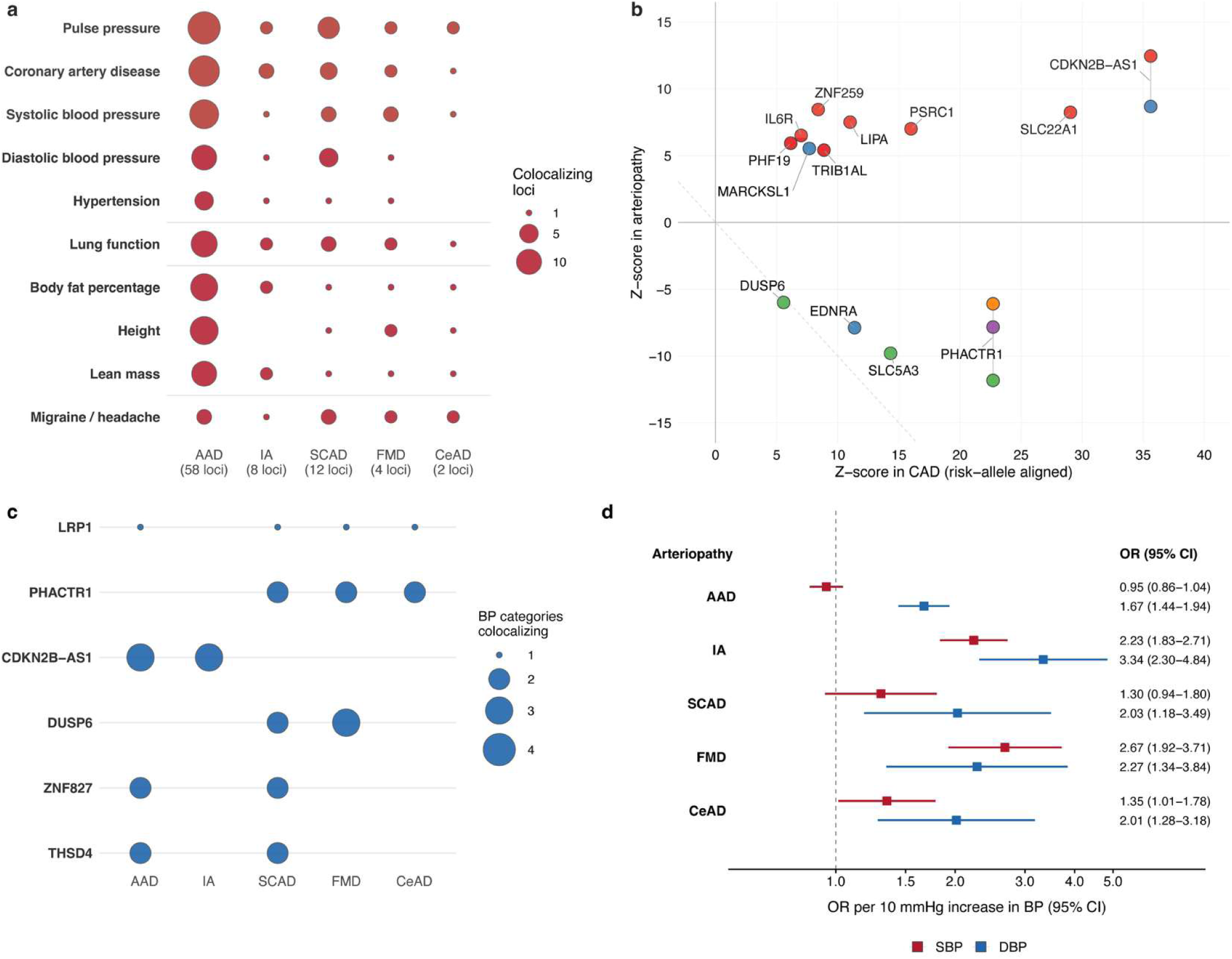
Shared phenotypes across arteriopathies in Genotype-Phenotype Map (a) Colocalization between arteriopathy GWAS and grouped categories of complex traits. Dots show the number of distinct loci colocalizing with at least one trait per group. (b) Effect directions at arteriopathy loci relative to CAD risk-increasing allele. Points above zero are concordant with CAD; below zero are discordant. Loci are coloured by arteriopathy: AAD (red), IA (blue), SCAD (green), FMD (purple), CeAD (orange) (c) Loci at which colocalizing associations with blood pressure-related phenotypes were observed in two or more arteriopathies, annotated by variant-to-gene mapping. Bubble size reflects the number of distinct blood pressure-related phenotypes colocalizing at each locus. (d) Two-sample Mendelian randomization estimates (IVW) for the effect of systolic (SBP) and diastolic (DBP) blood pressure on each arteriopathy, per 10 mmHg increase in BP. AAD, aortic aneurysm; CAD, coronary artery disease; CeAD, cervical artery dissection; FMD, fibromuscular dysplasia; IA, intracranial aneurysm; SCAD, spontaneous coronary artery dissection.

To assess whether arteriopathy-associated variants shared direction of effect with CAD, we compared z-scores at colocalizing loci reaching genome-wide significance in both the arteriopathy GWAS and CAD, after aligning effects to the CAD risk-increasing allele (Figure 4b). All eight loci associated with both AAD and CAD showed the same direction of effect, whereas loci associated with both CAD and SCAD, FMD, or CeAD showed the opposite direction of effect. *PHACTR1* was discordant in SCAD, FMD, and CeAD, *SLC5A3* and *DUSP6* were discordant in SCAD, and *EDNRA* was discordant in IA. IA was the only arteriopathy to show both patterns, with *CDKN2B-AS1* and *MARCKSL1* concordant with CAD and *EDNRA* discordant.

Blood pressure-related traits colocalized at multiple distinct loci within each arteriopathy (Table S16), with six loci shared across at least two arteriopathies (Figure 4c), including *LRP1* and *PHACTR1*. Given the colocalization between arteriopathy-associated loci and blood pressure-related traits, we next performed MR to assess whether blood pressure is causally associated with each arteriopathy. Among the blood pressure traits examined, genetically predicted diastolic blood pressure showed the most consistent association, with increased risk across all five arteriopathies (Figure 4d; Table S17).

### Proteome wide-Mendelian randomization

We next estimated the causal effects of protein abundance on arteriopathies using two-sample MR. After instrument selection and harmonization with outcome association data, 848–1683 cis-pQTLs for Olink-measured proteins (UK Biobank) and 910–1611 cis-pQTLs for SomaLogic-measured proteins (deCODE) were available for two-sample proteome-wide Mendelian randomization analyses, ranging from IA (fewest) to AAD (most) across both platforms (Table S18).

After correcting for multiple testing (FDR < 0.05), we identified 83 significant pQTL–outcome associations involving 67 unique proteins (Figure 5a; Table S19). Of these, 17 demonstrated support for a shared causal variant through colocalization analysis (PP.H4 ≥ 0.8), with an additional 3 demonstrating suggestive evidence (PP.H4 0.7–0.8). Colocalized pQTL associations included 1 protein for CeAD (ECM1), 1 for FMD (LIMA1), 2 for SCAD (ECM1, SPON1), 13 for AAD (including LTBP4, PCSK9, IL6R, LRP4, ECM1, and IL1RN; Table S19) and none for IA. ECM1 showed colocalized associations across two outcomes (CeAD, and AAD), representing the strongest cross-phenotype pQTL signal. SPON1 demonstrated consistent colocalized associations with SCAD across both Olink and SomaLogic platforms, with concordant effect directions.

**Figure 5.**
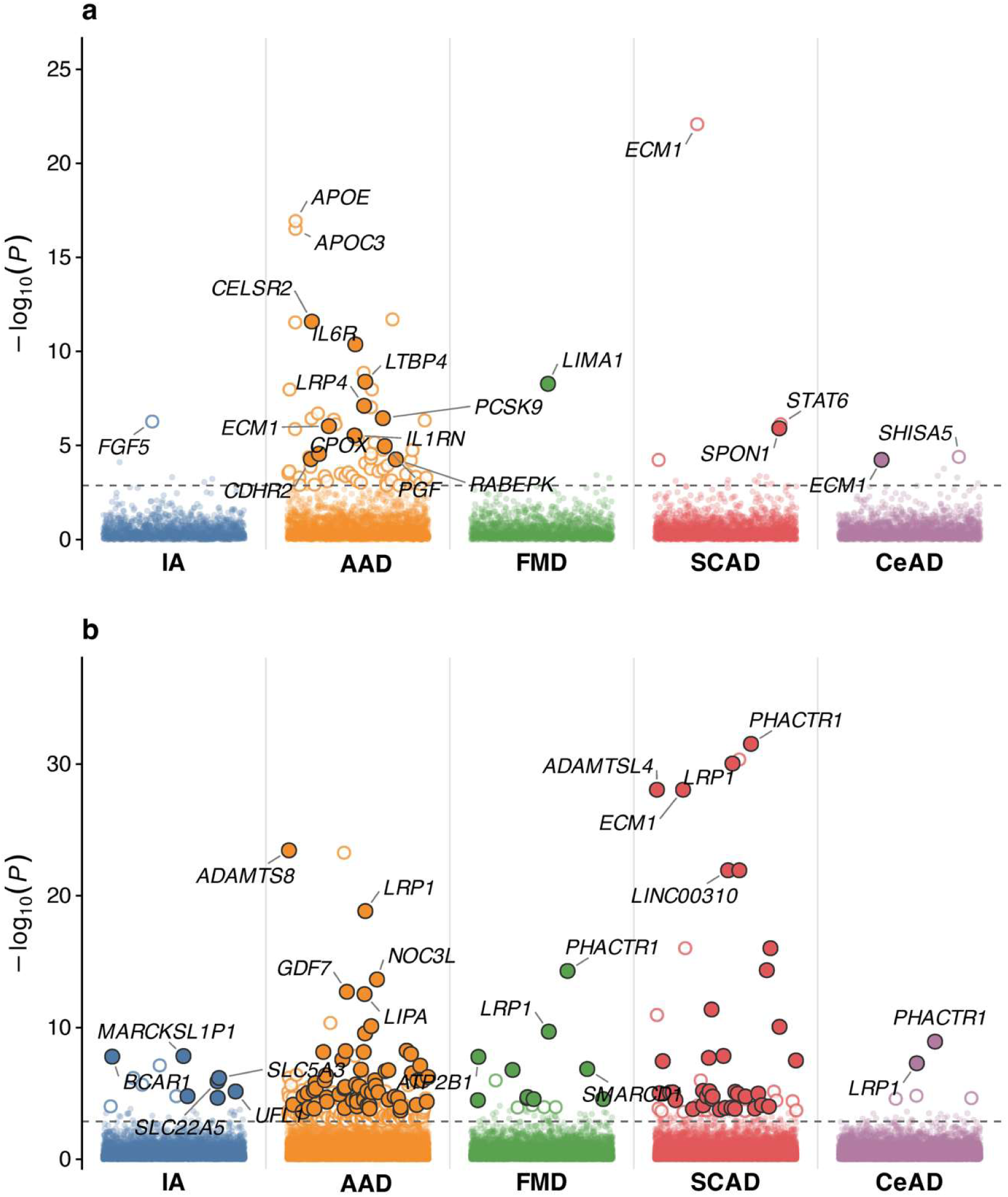
Cross-modality evidence for shared genes across arteriopathies. (a) Proteome-wide MR results across five arteriopathies in UKB and deCODE. Each point represents a gene–trait association, coloured by trait. Filled circles denote colocalization-validated associations (PP.H4 ≥ 0.8). Dashed line indicates FDR *P* = 0.05. (b) Transcriptome-wide MR results (arterial tissues; GTEx v10), coloured by trait. Filled and hollow circles as in (a).

To replicate proteome-wide MR findings, we constructed independent cis-pQTL instruments from the Fenland study (SomaScan v4, n = 10,708) and the SCALLOP consortium (Olink CVD-I, n = 30,931). Of 29 testable protein–trait associations, 25 (90%) replicated with concordant effect directions, including ECM1 and SPON1 for SCAD (Table S20).

### Transcriptome wide-Mendelian randomization

To complement our proteomic findings, we performed transcriptome-wide MR using cis-eQTLs from GTEx v10 across arterially-relevant tissues (aorta, tibial artery, coronary artery), fibroblasts and whole blood.

After FDR correction, we identified 760 significant eQTL–outcome associations (Figure 5b; Table S21). Of these, 715 underwent colocalization analysis, with 189 (26%) demonstrating strong evidence of a shared causal variant (PP.H4 ≥ 0.8) and 94 (13%) showing suggestive evidence (PP.H4 0.7–0.8; Table S21). 12 genes demonstrated colocalized eQTL associations across two or more outcomes, including *LRP1* (AAD, SCAD, FMD and CeAD), *PHACTR1* (SCAD FMD and CeAD), *FLNB* (AAD and SCAD) and *SLC24A3* (IA and SCAD). *MRPS6* and *SLC5A3* colocalized for both IA and SCAD but with opposing effect directions.

Cross-modality comparison of FDR-significant pQTL and eQTL MR results identified 29 overlapping gene–trait–tissue combinations for Olink (18 unique gene–trait pairs) and 13 for SomaScan (10 unique gene–trait pairs). Directional concordance between protein and expression MR estimates was 69.0% (20/29) for Olink and 46.2% (6/13) for SomaScan, with arterial tissues showing the highest agreement. Genes with opposing effect directions between pQTL and eQTL instruments included *ECM1* (AAD and SCAD) and *IL6R* (AAD).

### Functional analysis

Pathway enrichment of pQTL-MR significant proteins (n=67) identified extracellular matrix organization, collagen metabolism and TGF-β/SMAD signaling (Table S22). Arterial eQTL-MR significant genes (n=300) were enriched for smooth muscle cell differentiation and proliferation and vascular development (Table S23). Trait-specific enrichment results are provided in the Supplementary Material.

### Convergent cross-trait evidence

Twenty-eight genes showed genome-wide significant associations with at least three arteriopathies (Table S24). *CDKN2B-AS1* at the 9p21 locus was associated with all five traits, with strong pairwise colocalization supporting shared causal variants across IA, AAD and FMD. Ten genes were associated with four arteriopathies, including *LRP1*, *ECM1*, *ITGA1* and *ZNF827*. *ECM1* showed convergent MR and colocalization evidence across GWAS, pQTL and eQTL analyses for SCAD.

### Drug target assessment

Of 344 implicated genes across all analyses, 279 (81%) showed evidence of tractability by at least one therapeutic modality (109 small molecule, 170 antibody), and 23 corresponded to targets of approved drugs (Table S25–26). Several targets are already addressed by therapies with established cardiovascular applications, including *EDNRA* (endothelin receptor antagonists), *PCSK9* (PCSK9 inhibitors), and *GUCY1B1* (soluble guanylate cyclase stimulators). Other high-priority genes, including *ECM1* and *ITGA1*, showed predicted tractability despite lacking approved therapies.

## Discussion

In this study, we systematically evaluated the shared polygenic architecture across five arteriopathies. We demonstrated shared genetic liability across traits, identified 37 pleiotropic loci, and found evidence for shared causal variants at 13 loci across 24 trait pairs. We further found that blood pressure-related phenotypes were prominent among traits colocalizing with arteriopathy loci, and that many arteriopathy-associated variants showed effect directions opposite to those observed for CAD. Integrating transcriptome-and proteome-wide MR with colocalization, we identified 129 genes with evidence for causal roles in individual arteriopathies. Taken together, these analyses highlight both pleiotropic and trait-specific loci, and nominate relevant biological pathways and potential therapeutic targets.

Using complementary approaches, we show that arteriopathies share substantial heritable risk, with the strongest genetic overlap observed for SCAD–CeAD and CeAD–AAD. Subset-based meta-analysis identified 37 pleiotropic loci with evidence of multi-trait association, including *LRP1* and *PHACTR1*/*EDN1*, and further discussion of these previously well-characterized loci is provided in the Supplementary Discussion. 9p21 was identified as a shared locus across all five arteriopathies, extending previous studies that have implicated this region in IA and AAD.^31^ The 9p21 locus encompasses ANRIL, a long non-coding RNA that epigenetically represses *CDKN2A* and *CDKN2B* through recruitment of polycomb repressive complexes. This association was detected without corresponding pQTL or eQTL MR signals, which is consistent with evidence that 9p21 risk variants act through non-coding RNA mechanisms independent of mRNA or protein abundance.^32^

The *ECM1/ADAMTSL4* locus showed the strongest multi-modal convergence in our analyses, with both genes encoding secreted extracellular matrix proteins. ECM1 is a glycoprotein involved in basement membrane organization and angiogenesis, whereas ADAMTSL4 promotes fibrillin-1 microfibril assembly. Loss-of-function mutations in *ECM1* are associated with lipoid proteinosis, characterized by widespread extracellular matrix deposition and vascular abnormalities,^33^ whereas *ADAMTSL4* mutations can result in isolated ectopia lentis, phenocopying fibrillin-associated disorders.^34^ Proteome-wide MR identified ECM1 as the strongest cross-phenotype pQTL signal, while transcriptome-wide MR demonstrated arterial eQTL evidence for both genes. Notably, genetically predicted *ECM1* effects were discordant across molecular layers: higher arterial expression was associated with lower risk of SCAD and AAD, whereas higher circulating ECM1 protein levels were associated with increased risk of SCAD, CeAD, and AAD. The circulating protein signal was driven by the same missense variant across both Olink and SomaLogic platforms, whereas the arterial eQTL was instrumented by a different variant. This suggests that tissue-level transcription and circulating protein abundance may reflect distinct regulatory mechanisms at this locus.

The 21q22 locus also emerged as a shared signal across all arteriopathies. This region contains two tightly co-regulated genes, *MRPS6* and *SLC5A3*, both of which are supported by arterial eQTL colocalization for SCAD and IA. However, our tiered annotation framework prioritized *MRPS6* as the most likely effector gene at this locus. This locus has been identified in GWAS of coronary artery disease, yet fine-mapping has been unable to distinguish between candidates.^35^ Our lead variant, rs28451064, is associated with increased arterial expression of both genes, and this tight co-regulation limits statistical resolution.^36^ Notably, rs28451064 also lies within credible sets for diastolic blood pressure, linking this locus to vascular tone and arterial compliance. *MRPS6* encodes a mitochondrial ribosomal protein involved in oxidative phosphorylation, and its perturbation induces mitochondrial stress and apoptosis,^37^ changes that may impair vascular smooth muscle function, which depends on mitochondrial ATP production.^38^ Together, these observations suggest that 21q22 may represent a shared regulatory locus influencing vascular integrity via effects on mitochondrial energy metabolism.

Several of the ten loci associated with four or more arteriopathies converge on pathways related to the function of vascular smooth muscle cells, regulation of extracellular matrix, and arterial wall integrity. Among these, *ZNF827* has been functionally implicated as a regulator of gene expression in vascular SMCs and fibroblasts, supporting a plausible mechanistic link to susceptibility to arteriopathy.^39^ Similarly, *ITGA1*, encodes the α1 subunit of the α1β1 integrin, a collagen and laminin receptor on vascular smooth muscle cells, that regulates adhesion, migration, and extracellular matrix remodeling.^40^ Experimental studies show that α1β1 is upregulated during neointima formation after vascular injury, and that loss of α1β1 or β1 integrin impairs matrix homeostasis and vascular wall stability.^41,42^ Our findings extend previous genetic evidence implicating *ITGA1* in SCAD and suggest that common variation at this locus, potentially acting through altered α1β1 signaling and mechanotransduction at the smooth muscle cell-collagen interface, contributes more broadly to susceptibility across multiple arteriopathies. Although direct therapeutic targeting of *ITGA1* has not yet been established, the broader druggability of integrin receptors supports the translational plausibility of this signal.

*SLC22A5*, encoding the high-affinity carnitine transporter OCTN2, was implicated at 5q31 across four arteriopathies and was further supported by strong arterial eQTL colocalization. OCTN2 mediates cellular carnitine uptake required for mitochondrial fatty-acid β-oxidation, and impaired function can lead to systemic carnitine deficiency with cardiomyopathy. Disrupted carnitine homeostasis impairs mitochondrial function and alters nitric oxide signaling in endothelial models,^43^ and more broadly, metabolic reprogramming including fatty-acid oxidation is a recognized feature of vascular smooth muscle cell phenotypic switching.^44^ Notably, the broader 5q31 block contains additional candidates, including *P4HA2*, which has been functionally implicated in the progression of IA,^45^ and further work will be needed to resolve the effector gene at 5q31.

Multi-trait genome-wide association analysis substantially increased discovery power, with seven trait-specific associations across six loci reaching genome-wide significance only through multi-trait analysis. Of these, three associations at two loci had not been previously reported for their respective traits, including one for CeAD and SCAD, prioritized to *SLC39A8*, and one for IA prioritized to *FGF5*, which has recently been reported in a large GWAS meta-analysis but has not yet been catalogued. The missense variant rs13107325 (A391T) is a highly pleiotropic variant at the *SLC39A8* locus, with established associations with blood pressure, cardiometabolic traits, and autoimmune diseases.^46^ ZIP8 (encoded by *SLC39A8*) is a divalent metal cation transporter essential for zinc and manganese homeostasis.^47^ Functional studies demonstrate that ZIP8 regulates extracellular matrix turnover through the zinc-MTF1-ADAMTS axis. Slc39a8-null mice exhibit cardiac extracellular matrix accumulation and reduced expression of ADAMTS metalloproteinases, while ZIP8 knockdown in endothelial cells decreases *ADAMTS1* transcription through attenuated MTF1 activity.^47^ These results suggest that metal ion dyshomeostasis may compromise arterial wall integrity by impairing ECM remodeling, providing a mechanistic link between this pleiotropic variant and susceptibility to CeAD and SCAD.

Headache-and migraine-related phenotypes emerged as recurrent colocalizing traits across arteriopathy-associated loci in GPMap. Prior studies have shown substantial shared genetic architecture between migraine and CeAD, including overlap at *PHACTR1*/*EDN1*, *LRP1* and *ECM1*.^48^ Migraine has also found to be associated with other arteriopathies, including SCAD, FMD and IA.^11,49,50^ Our findings extend this picture by suggesting that shared genetic signals between arteriopathies and headache-and migraine-related phenotypes are present across the broader arteriopathy spectrum at multiple independent loci. Blood pressure-related phenotypes were also prominent in GPMap, with colocalizing associations observed across multiple loci in each arteriopathy. Only six loci showed overlap in at least two arteriopathies, all of which had also been identified in ASSET. MR further supported a causal relationship between blood pressure and risk of arteriopathy, with diastolic blood pressure associated with all five traits.

Our findings extend prior observations of the inverse genetic relationship between SCAD and CAD to a broader pattern across the arteriopathies. Whereas all loci shared between AAD and CAD were directionally concordant, loci shared between CAD and SCAD, FMD and CeAD showed opposite directions of effect, with IA showing both patterns. Although opposing variant effects have previously been described at the *PHACTR1/EDN1* locus and in SCAD more broadly,^12^ our results suggest that this is not confined to a single locus or arteriopathy, but may instead reflect broader differences in how these loci influence arterial wall remodeling and structural integrity compared with the mechanisms underlying atherosclerotic CAD.

Several pleiotropic loci identified in our analyses map to pathways of therapeutic interest. Among these, *EDNRA* is particularly notable because it encodes the endothelin A receptor, a central mediator of vasoconstriction and vascular remodeling. Although the association at this locus was limited to IA and SCAD, it provides one of the clearest translational links in our study, given that endothelin receptor antagonists are already approved for pulmonary arterial hypertension and, more recently, have shown efficacy in resistant hypertension. These observations support endothelin signaling as a tractable pathway with potential relevance across arteriopathies, particularly in light of the broader overlap we observed with blood pressure-related phenotypes. Although *EDNRA* provides the clearest near-term translational link, additional loci such as *ITGA1*, *LRP1* and *ECM1* may be of longer-term interest, given emerging evidence for the tractability of integrin-mediated and receptor-targeted approaches.

This study has several important strengths. By leveraging cross-trait analytical frameworks we increased statistical power to detect shared genetic associations across arteriopathies, some of which remain individually underpowered. We integrated multiple complementary methods which provide convergent evidence across independent analytical approaches. To prioritize likely causal genes, we combined annotation pipelines with eQTL and pQTL colocalization, gene-based analyses, and manual curation, allowing more confident assignment of genes to associated loci. Finally, downstream biological, pathway, and druggability analyses enabled interpretation of genetic findings in a mechanistic and translational context.

Several limitations should be considered. Genome-wide association studies primarily capture common single-nucleotide variants and not rare variants, insertions and deletions, or structural variation, which may contribute to the risk of arteriopathy. Sample sizes for some traits was modest, and limited power to detect trait-specific associations. In addition, the AAD phenotype encompassed both abdominal and thoracic aortopathies, potentially introducing heterogeneity and attenuating associations specific to individual aortic subtypes, which have partly distinct biological underpinnings.

Proteomic analyses rely on aptamer-and antibody-based platforms, which may not fully reflect circulating or tissue-level protein abundance, and available eQTL and pQTL datasets are derived largely from healthy volunteers with limited sample sizes. Our analyses were restricted to individuals of European ancestry, which may limit generalizability to other populations. Finally, while genetic and integrative genomic approaches provide strong evidence for causal involvement, further functional studies will be required to resolve causal variants, define cell-type-specific mechanisms, and establish how shared and distinct genetic pathways contribute to arteriopathy risk.

In conclusion, our findings support shared genetic architecture across anatomically distinct arteriopathies, identify previously uncharacterized pleiotropic loci, and highlight convergence on blood pressure-related traits and extracellular matrix-related vascular pathways. Together, these analyses prioritize pleiotropic genes that may inform drug repurposing and the development of novel therapeutic strategies.

## Nonstandard Abbreviations and Acronyms

IA: Intracranial Aneurysm
AAD: Aortic Aneurysm and Dissection
SCAD: Spontaneous Coronary Artery Dissection
FMD: Fibromuscular Dysplasia
CAD: Coronary Artery Disease
MR: Mendelian Randomization
GWAS: Genome-wide Association Study
FDR: False Discovery Rate
MTAG: Multi-Trait Analysis of GWAS
GPMAP: Genotype-Phenotype Map

## References

1. Harris SL, Lindsay ME. Role of clinical genetic testing in the management of aortopathies. Current Cardiology Reports. 2021;23:10.

2. Meester JA, Verstraeten A, Schepers D, Alaerts M, Van Laer L, Loeys BL. Differences in manifestations of Marfan syndrome, Ehlers-Danlos syndrome, and Loeys-Dietz syndrome. Annals of cardiothoracic surgery. 2017;6:582.

3. Weerakkody R, Ross D, Parry DA, Ziganshin B, Vandrovcova J, Gampawar P, Abdullah A, Biggs J, Dumfarth J, Ibrahim Y, et al. Targeted genetic analysis in a large cohort of familial and sporadic cases of aneurysm or dissection of the thoracic aorta. Genetics in Medicine. 2018;20:1414–1422. doi: 10.1038/gim.2018.27

4. Kim ESH, Saw J, Kadian-Dodov D, Wood M, Ganesh SK. FMD and SCAD: Sex-Biased Arterial Diseases With Clinical and Genetic Pleiotropy. Circulation Research. 2021;128:1958–1972. doi: 10.1161/CIRCRESAHA.121.318300

5. Vlak MHM, Algra A, Brandenburg R, Rinkel GJE. Prevalence of unruptured intracranial aneurysms, with emphasis on sex, age, comorbidity, country, and time period: a systematic review and meta-analysis. The Lancet Neurology. 2011;10:626–636. doi: 10.1016/S1474-4422(11)70109-0

6. Le Grand Q, Ecker Ferreira L, Metso TM, Schilling S, Tatlisumak T, Grond-Ginsbach C, Engelter ST, Lyrer P, Majersik JJ, Worrall BB. Genetic insights on the relation of vascular risk factors and cervical artery dissection. Journal of the American College of Cardiology. 2023;82:1411–1423.

7. Liang JJ, Prasad M, Tweet MS, Hayes SN, Gulati R, Breen JF, Leng S, Vrtiska TJ. A novel application of CT angiography to detect extracoronary vascular abnormalities in patients with spontaneous coronary artery dissection. Journal of cardiovascular computed tomography. 2014;8:189–197.

8. Keser Z, Chiang CC, Benson JC, Pezzini A, Lanzino G. Cervical Artery Dissections: Etiopathogenesis and Management. Vasc Health Risk Manag. 2022;18:685–700. doi: 10.2147/vhrm.S362844

9. Southerland AM, Meschia JF, Worrall BB. Shared associations of nonatherosclerotic, large-vessel, cerebrovascular arteriopathies: considering intracranial aneurysms, cervical artery dissection, moyamoya disease and fibromuscular dysplasia. Current opinion in neurology. 2013;26:13–28.

10. Saw J, Ricci D, Starovoytov A, Fox R, Buller CE. Spontaneous Coronary Artery Dissection: Prevalence of Predisposing Conditions Including Fibromuscular Dysplasia in a Tertiary Center Cohort. JACC: Cardiovascular Interventions. 2013;6:44–52. doi: 10.1016/j.jcin.2012.08.017

11. Bonacina S, Grassi M, Zedde M, Zini A, Bersano A, Gandolfo C, Silvestrelli G, Baracchini C, Cerrato P, Lodigiani C. Clinical features of patients with cervical artery dissection and fibromuscular dysplasia. Stroke. 2021;52:821–829.

12. Adlam D, Berrandou T-E, Georges A, Nelson CP, Giannoulatou E, Henry J, Ma L, Blencowe M, Turley TN, Yang M-L. Genome-wide association meta-analysis of spontaneous coronary artery dissection identifies risk variants and genes related to artery integrity and tissue-mediated coagulation. Nature genetics. 2023;55:964–972.

13. Debette S, Kamatani Y, Metso TM, Kloss M, Chauhan G, Engelter ST, Pezzini A, Thijs V, Markus HS, Dichgans M. Common variation in PHACTR1 is associated with susceptibility to cervical artery dissection. Nature genetics. 2015;47:78–83.

14. Georges A, Yang M-L, Berrandou T-E, Bakker MK, Dikilitas O, Kiando SR, Ma L, Satterfield BA, Sengupta S, Yu M, et al. Genetic investigation of fibromuscular dysplasia identifies risk loci and shared genetics with common cardiovascular diseases. Nature Communications. 2021;12:6031. doi: 10.1038/s41467-021-26174-2

15. Bakker MK, van der Spek RA, van Rheenen W, Morel S, Bourcier R, Hostettler IC, Alg VS, van Eijk KR, Koido M, Akiyama M. Genome-wide association study of intracranial aneurysms identifies 17 risk loci and genetic overlap with clinical risk factors. Nature genetics. 2020;52:1303–1313.

16. Verma A, Huffman JE, Rodriguez A, Conery M, Liu M, Ho Y-L, Kim Y, Heise DA, Guare L, Panickan VA. Diversity and scale: Genetic architecture of 2068 traits in the VA Million Veteran Program. Science. 2024;385:eadj1182.

17. Bulik-Sullivan BK, Loh P-R, Finucane HK, Ripke S, Yang J, Patterson N, Daly MJ, Price AL, Neale BM. LD Score regression distinguishes confounding from polygenicity in genome-wide association studies. Nature genetics. 2015;47:291–295.

18. Bhattacharjee S, Rajaraman P, Jacobs KB, Wheeler WA, Melin BS, Hartge P, Yeager M, Chung CC, Chanock SJ, Chatterjee N. A subset-based approach improves power and interpretation for the combined analysis of genetic association studies of heterogeneous traits. The American Journal of Human Genetics. 2012;90:821–835.

19. Zhu X, Feng T, Tayo BO, Liang J, Young JH, Franceschini N, Smith JA, Yanek LR, Sun YV, Edwards TL. Meta-analysis of correlated traits via summary statistics from GWASs with an application in hypertension. The American Journal of Human Genetics. 2015;96:21–36.

20. Turley P, Walters RK, Maghzian O, Okbay A, Lee JJ, Fontana MA, Nguyen-Viet TA, Wedow R, Zacher M, Furlotte NA. Multi-trait analysis of genome-wide association summary statistics using MTAG. Nature genetics. 2018;50:229–237.

21. Wallace C. A more accurate method for colocalisation analysis allowing for multiple causal variants. PLOS Genetics. 2021;17:e1009440. doi: 10.1371/journal.pgen.1009440

22. Elmore AR, Hanson AL, Leyden GM, Johnson J, Davey Smith G, Paternoster L, Gaunt TR, Hemani G. Building The Human Genotype-Phenotype Map to Harness Pleiotropy and Refine Disease Mechanisms. medRxiv. 2026:2026.2002.2019.26346618. doi: 10.64898/2026.02.19.26346618

23. Aragam KG, Jiang T, Goel A, Kanoni S, Wolford BN, Atri DS, Weeks EM, Wang M, Hindy G, Zhou W, et al. Discovery and systematic characterization of risk variants and genes for coronary artery disease in over a million participants. Nat Genet. 2022;54:1803–1815. doi: 10.1038/s41588-022-01233-6

24. Sun BB, Chiou J, Traylor M, Benner C, Hsu Y-H, Richardson TG, Surendran P, Mahajan A, Robins C, Vasquez-Grinnell SG. Plasma proteomic associations with genetics and health in the UK Biobank. Nature. 2023;622:329–338.

25. Ferkingstad E, Sulem P, Atlason BA, Sveinbjornsson G, Magnusson MI, Styrmisdottir EL, Gunnarsdottir K, Helgason A, Oddsson A, Halldorsson BV. Large-scale integration of the plasma proteome with genetics and disease. Nature genetics. 2021;53:1712–1721.

26. Folkersen L, Gustafsson S, Wang Q, Hansen DH, Hedman ÅK, Schork A, Page K, Zhernakova DV, Wu Y, Peters J, et al. Genomic and drug target evaluation of 90 cardiovascular proteins in 30,931 individuals. Nature Metabolism. 2020;2:1135–1148. doi: 10.1038/s42255-020-00287-2

27. Pietzner M, Wheeler E, Carrasco-Zanini J, Cortes A, Koprulu M, Wörheide MA, Oerton E, Cook J, Stewart ID, Kerrison ND, et al. Mapping the proteo-genomic convergence of human diseases. Science. 2021;374:eabj1541. doi: doi:10.1126/science.abj1541

28. Consortium G. The GTEx Consortium atlas of genetic regulatory effects across human tissues. Science. 2020;369:1318–1330.

29. Tang H, Walker VM, Gaunt TR. Evaluating the causal relationships between urate, blood pressure, and kidney function in the general population: a two-sample Mendelian Randomization study. medRxiv. 2024:2024.2004.2025.24306305. doi: 10.1101/2024.04.25.24306305

30. Watanabe K, Taskesen E, Van Bochoven A, Posthuma D. Functional mapping and annotation of genetic associations with FUMA. Nature communications. 2017;8:1826.

31. Helgadottir A, Thorleifsson G, Magnusson KP, Grétarsdottir S, Steinthorsdottir V, Manolescu A, Jones GT, Rinkel GJ, Blankensteijn JD, Ronkainen A. The same sequence variant on 9p21 associates with myocardial infarction, abdominal aortic aneurysm and intracranial aneurysm. Nature genetics. 2008;40:217–224.

32. Holdt LM, Stahringer A, Sass K, Pichler G, Kulak NA, Wilfert W, Kohlmaier A, Herbst A, Northoff BH, Nicolaou A, et al. Circular non-coding RNA ANRIL modulates ribosomal RNA maturation and atherosclerosis in humans. Nature Communications. 2016;7:12429. doi: 10.1038/ncomms12429

33. Hamada T, McLean WI, Ramsay M, Ashton GH, Nanda A, Jenkins T, Edelstein I, South AP, Bleck O, Wessagowit V. Lipoid proteinosis maps to 1q21 and is caused by mutations in the extracellular matrix protein 1 gene (ECM1). Human molecular genetics. 2002;11:833–840.

34. Hubmacher D, Apte SS. ADAMTS proteins as modulators of microfibril formation and function. Matrix Biology. 2015;47:34–43.

35. van der Harst P, Verweij N. Identification of 64 Novel Genetic Loci Provides an Expanded View on the Genetic Architecture of Coronary Artery Disease. Circulation Research. 2018;122:433–443. doi: doi:10.1161/CIRCRESAHA.117.312086

36. Beaney KE, Smith AJP, Folkersen L, Palmen J, Wannamethee SG, Jefferis BJ, Whincup P, Gaunt TR, Casas JP, Ben-Shlomo Y, et al. Functional Analysis of the Coronary Heart Disease Risk Locus on Chromosome 21q22. Disease Markers. 2017;2017:1096916. doi: 10.1155/2017/1096916

37. Lin D, Yu J, Lin L, Ou Q, Quan H. MRPS6 modulates glucose-stimulated insulin secretion in mouse islet cells through mitochondrial unfolded protein response. Scientific Reports. 2023;13:16173. doi: 10.1038/s41598-023-43438-7

38. Liu Y-F, Zhu J-J, Yu Tian X, Liu H, Zhang T, Zhang Y-P, Xie S-A, Zheng M, Kong W, Yao W-J, et al. Hypermethylation of mitochondrial DNA in vascular smooth muscle cells impairs cell contractility. Cell Death & Disease. 2020;11:35. doi: 10.1038/s41419-020-2240-7

39. Liu Y, Liu L, Esmael A, Fustier M-A, Georges A, Bouatia-Naji N. <em>ZNF827</em> pleiotropic cardiovascular risk locus involves regulation by Nuclear factor-1. bioRxiv. 2024:2024.2001.2023.576845. doi: 10.1101/2024.01.23.576845

40. Obata H, Hayashi K, Nishida W, Momiyama T, Uchida A, Ochi T, Sobue K. Smooth muscle cell phenotype-dependent transcriptional regulation of the alpha1 integrin gene. J Biol Chem. 1997;272:26643–26651. doi: 10.1074/jbc.272.42.26643

41. Abraham S, Kogata N, Fässler R, Adams RH. Integrin β1 Subunit Controls Mural Cell Adhesion, Spreading, and Blood Vessel Wall Stability. Circulation Research. 2008;102:562–570. doi: 10.1161/CIRCRESAHA.107.167908

42. Turlo KA, Scapa J, Bagher P, Jones AW, Feil R, Korthuis RJ, Segal SS, Iruela-Arispe ML. β1-integrin is essential for vasoregulation and smooth muscle survival in vivo. Arterioscler Thromb Vasc Biol. 2013;33:2325–2335. doi: 10.1161/atvbaha.112.300648

43. Sharma S, Black SM. CARNITINE HOMEOSTASIS, MITOCHONDRIAL FUNCTION, AND CARDIOVASCULAR DISEASE. Drug Discov Today Dis Mech. 2009;6:e31–e39. doi: 10.1016/j.ddmec.2009.02.001

44. Shi J, Yang Y, Cheng A, Xu G, He F. Metabolism of vascular smooth muscle cells in vascular diseases. American Journal of Physiology-Heart and Circulatory Physiology. 2020;319:H613–H631. doi: 10.1152/ajpheart.00220.2020

45. Li L, Wang J, Ren S, Hao X. P4HA2 knockdown prevents the progression of intracranial aneurysm by inducing prolyl hydroxylation of YAP1. Neurosurgical Review. 2024;47:858. doi: 10.1007/s10143-024-03101-9

46. Pickrell JK, Berisa T, Liu JZ, Ségurel L, Tung JY, Hinds DA. Detection and interpretation of shared genetic influences on 42 human traits. Nature Genetics. 2016;48:709–717. doi: 10.1038/ng.3570

47. Lin W, Li D, Cheng L, Li L, Liu F, Hand NJ, Epstein JA, Rader DJ. Zinc transporter Slc39a8 is essential for cardiac ventricular compaction. The Journal of clinical investigation. 2018;128:826–833.

48. Daghlas I, Sargurupremraj M, Danning R, Gormley P, Malik R, Amouyel P, Metso T, Pezzini A, Kurth T, Debette S, et al. Migraine, Stroke, and Cervical Arterial Dissection. Neurology Genetics. 2022;8:00. doi: doi:10.1212/NXG.0000000000000653

49. Kok SN, Hayes SN, Cutrer FM, Raphael CE, Gulati R, Best PJM, Tweet MS. Prevalence and Clinical Factors of Migraine in Patients With Spontaneous Coronary Artery Dissection. Journal of the American Heart Association. 2018;7:e010140. doi: doi:10.1161/JAHA.118.010140

50. Daghlas I, Rist PM, Chasman DI. Genetically proxied liability to migraine and risk of intracranial aneurysm and subarachnoid hemorrhage. Headache: The Journal of Head and Face Pain. 2025;65:391–398.

